# Serum protein and imaging biomarkers after intermittent steroid treatment in muscular dystrophy

**DOI:** 10.1101/2024.06.14.24308858

**Authors:** Alexander B. Willis, Aaron S. Zelikovich, Robert Sufit, Senda Ajroud-Driss, Krista Vandenborne, Alexis R. Demonbreun, Abhinandan Batra, Glenn A. Walter, Elizabeth M. McNally

## Abstract

**Background:** Weekly Steroids in Muscular Dystrophy (WSiMD) was a pilot study to evaluate once weekly prednisone in patients with Limb Girdle and Becker muscular dystrophy (LGMD and BMD, respectively). At study endpoint, there were trends towards increased lean mass, reduced fat mass, reduced creatine kinase and improved motor function. The investigation was motivated by studies in mouse muscular dystrophy models in which once weekly glucocorticoid exposure enhanced muscle strength and reduced fibrosis.

**Methods:** WSiMD participants provided blood samples for aptamer serum profiling at baseline and after 6 months of weekly steroids. A subset completed magnetic resonance (MR) evaluation of muscle at study onset and endpoint.

**Results/Conclusions:** At baseline compared to age and sex-matched healthy controls, the aggregate serum protein profile in the WSiMD cohort was dominated by muscle proteins, reflecting leak of muscle proteins into serum. Disease status produced more proteins differentially present in serum compared to steroid-treatment effect. Nonetheless, a response to prednisone was discernable in the WSiMD cohort, even at this low dose. Glucocorticoids downregulated muscle proteins and upregulated certain immune process- and matrix-associated proteins. Muscle MR fat fraction showed trends with functional status. The prednisone-responsive markers could be used in larger trial of prednisone efficacy.

## INTRODUCTION

Glucocorticoid steroids are used to treat Duchenne Muscular Dystrophy, where their use has been shown to prolong ambulatory duration and to correlate with improved cardiac and respiratory muscle function ^1,2^. The most commonly used glucocorticoid agents in DMD are prednisone and deflazacort, typically across a range of dosing regimens including once daily, high dose weekend, and “ten day on/off” strategies ^3^. With DMD patients having longer lifespans, in part due to steroid use, non-invasive respiratory support, and supportive cardiac care, many DMD patients are now receiving glucocorticoids for two or more decades.

Prolonged use of glucocorticoids in DMD associates with adverse outcomes like delayed puberty with altered testosterone levels, adrenal suppression, obesity, osteoporosis, cataracts and metabolic syndrome ^4,5^. Among the more troubling side effects of steroid use in DMD are obesity and behavioral issues, which can lead to using alternative dosing strategies or cessation of steroid use. High-dose steroid given twice weekly on weekends has been found to have benefit in DMD ^6^.

In an effort to identify glucocorticoid dosing that provided benefit with fewer side effects, we evaluated the use of once weekly steroids in mouse models of DMD and LGMD ^7,8^. We identified that once weekly steroids improved muscle strength and reduced fibrosis in the *mdx* model of DMD as well as in mouse models of LGMD due to sarcoglycan or dysferlin mutations, *Sgcg* and *Dysf*, respectively ^7,8^. In the *mdx* model, where the molecular consequences of this intermittent steroid dosing were detailed, once weekly steroids “reprogrammed” muscle, seen as altered gene expression accompanied by epigenetic marks consistent with the gene expression changes ^9^. Notably, the KLF15 and MEF2C pathways were specifically upregulated by once weekly steroids, and the gene expression cascades driven by the transcriptional regulators KLF15 and MEF2C altered nutrient uptake into muscle ^7,9^. Accompanying these muscle effects was a reduction in inflammatory pathways and an ability to recover better from muscle injury. We found that the effect of glucocorticoid dosing was dependent on time of day dosing ^10^. Furthermore, mice with genetically disrupted circadian rhythm no longer benefitted from once weekly dosing ^10^.

To evaluate whether this same steroid dosing strategy was effective in human muscular dystrophy (MD), we conducted a pilot, open label study in LGMD patients and one BMD patient. The Weekly Steroids in Muscular Dystrophy (WSiMD) study recruited participants for once weekly glucocorticoid dosed at 0.75 -1.0 mg/kg/week, depending on body mass, taken once weekly in the evening ^11^. Participants included a range of patients with MD subtypes and disease progression, including approximately one-third ambulatory, one-third nonambulatory and one-third transitioning to nonambulatory. All patients were adults, and males and females were included. Overall, we observed that once weekly steroid dosing was well tolerated with few side effects over six months. Participants showed trends towards improved lean body mass, reduced CK, and greater distance walked in 6 mintues ^11^.

Serum profiling and magnetic resonance imaging (MRI) and magnetic resonance spectroscopy (MRS) in DMD patients provide important biomarker correlates of disease trajectories and treatment effects ^12–16^. The goal of this study was to identify clinically relevant biomarkers associated with the WSiMD cohort as an aggregate group, and to evaluate whether we could discern a biomarker signature of intermittent prednisone treatment at this comparatively low dose of glucocorticoid exposure. Past aptamer profiling studies in MD have focused on DMD, including steroid responsiveness, but these studies differ in their focus on children and males ^12,17^. We evaluated the serum aptamer profiles in the WSiMD cohort and compared to these prior studies to assess MD markers as a whole and to identify potentially useful steroid responsive biomarkers.

## METHODS

### Ethical Declaration

As previously noted, the WSiMD study was (NCT04054375, Date of Registration 13/08/2019 (Aug 13, 2019)) was approved by the Institutional Review Board at Northwestern University Feinberg School of Medicine (STU00208443) ^11^. All participants provided written informed consent (n=20) following a discussion of study goals, design, risks, and benefits preceding enrollment in compliance with the ethical standards of the Helsinki declaration of 1975.

### Inclusion and Exclusion Criteria

All WSiMD participants (n=20) were recruited from the Northwestern University Muscular Dystrophy Association clinic between June 2019 and January 2020. WSiMD participants were between 18-65 years (mean age of 35), electrocardiogram within 2 months prior to enrollment with no evidence of atrial fibrillation or recent myocardial infarction, genetic testing confirming BMD or LGMD variant, echocardiogram showing left ventricular ejection fraction greater than 25%, and no changes to medication within three months of enrollment. Exclusion criteria were a history of oral steroid use in the preceding three years for longer than one month, history of diabetes mellitus, BMI greater than 35 kg/m^2^, uncontrolled hypertension, congestive heart failure, chronic kidney disease, full time ventilator dependency, orthopedic surgery within past 6 months, current pregnancy, and any history of tuberculosis.

### Study Design

Participants completed a 24-week, open-label trial. At week 0, baseline measurements included heart rate, blood pressure, weight, height, forced vital capacity, and muscle imaging. These same measures were repeated at study endpoint at 24 weeks. Whole blood samples were collected at week 0 and week 24. The Vignos and Brooke scales were used for lower and upper extremity measurements, and the NorthStar Ambulatory assessment for Dysferlinopathy (NSAD) was used. Each participant was tested for grip strength. Six-minute walk test and 10-meter run test data were collected on ambulatory patients. All data was collected on site by study coordinators and therapists. Study participants received monthly phone calls at weeks 4, 8, 12, 16, and 20 from study coordinators confirming medication adherence, dosage, and any adverse events. DEXA studies were completed at week 0 and study endpoint for most participants. Completion of all study data points was limited for some participants due to the COVID-19 pandemic in March 2020.

### Magnetic Resonance Imaging

Participants (n=13) completed upper and lower extremity imaging according to previously established methods at study onset and endpoint. Quadriceps and Triceps surae muscles were analyzed for fat fraction ^18–21^. Muscle fat fraction (FF) was assessed using both an imaging method (chemical shift encoded or Dixon MRI) and the gold standard of volume-localized magnetic resonance spectroscopy (MRS). Fat fraction images were calculated using a muscle-based multi-peak analysis^1^ from a 12-point Dixon dataset, which combined three multi-slice, multi-echo scans (TR=430ms, 18 four mm slices, 20 degree flip angle), each obtained with four unique echo times [TEs] with overall TEs of 2.4, 3.6, 4.8, 5.9, 8.4, 9.4, 9.6, 13.0, 13.2, 14.3, 17. 19.1 ms. Single voxel ^1^H-MRS muscle FF was derived from Vastus Lateralis and Soleus muscle respectively in both the lower leg and thigh using Stimulated Echo Acquisition Mode (STEAM;TR= 9,000ms, TM=16ms, BW=2KHz, complex data points= 2,048) without water suppression and with multiple echo times (TE= 11, 27, 54, and 243 ms) to measure water T_2_. ^1^H MRS spectra were corrected for T_2_ relaxation as previously described, and the water and lipid regions were integrated to determine muscle FF ^2^. In cases where disease progression eliminated any visible VL and Sol muscle an alternative upper and/or lower limb muscle was selected. Cross-validation of the imaging sequence and system quality control were conducted at each imaging session using a co-axial phantom^3^ and by co-registering MRI and MRS-determined FF ^1^.

To quantify FF based on Dixon imaging, custom-written IDL software was used to generate pixel by pixel FF maps from water and fat images of both LL and thigh. Regions of interest (ROIs) were manually drawn on the FF maps for the four muscles combining to form quadriceps (Vastus lateralis (VL), Vastus intermedius (VI), Vastus medius (VM), Rectus femoris (RF)) and three lower leg muscles combining to form Triceps surae (soleus (Sol), medial gastrocnemius (MG), lateral gastrocnemius (LG)).Each muscle was analyzed on the same three consecutive slices based on pre-determined landmarks. The mean FF for each slice was determined for all pixels within ROI using the following formulae:

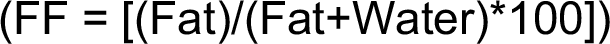

Muscle fat fraction for the four thigh and the three LL muscles were averaged to get average fat fraction for quadriceps and Triceps surae muscle groups.

### Serum Aptamer Profiling and Analysis

Sera was collected from WSiMD participants at study onset and after 24 weeks of once weekly prednisone. Age/sex matched healthy individuals without steroid exposure were used for controls. Sera was analyzed using SomaScan aptamer profiling. The SomaScan platform utilizes modified DNA aptamers to detect 6411 unique proteins from small volumes of serum, with a subset of proteins containing multiple aptamers for a total of 7322 aptamers. Analysis was conducted by importing data into R using the SomalogicsIO R package, and subsequent analysis was performed in R Studio. Serum biomarkers associating with disease and prednisone treatment were evaluated using a linear mixed model from the variancePartition R package, with a model of y ∼ Disease + Prednisone + (1|Subject), with a random affect accounting for patient to account for basal differences in the serum profile between patients. Disease and prednisone-responsive terms were then extracted using the TopTable function, and adjusted p value and logFC thresholds were used as described in the results. Pathway analysis was performed using the ClusterProfiler package on significantly upregulated or downregulated terms. Lists of significantly affected terms between disease and treatment were compared to determine the set of disease-associated treatment-responsive biomarkers.

Spearman correlation was used to assess the relationships between age, muscle functional test outcomes, fat fraction and water T_2_ MRI/MRs measures, and DEXA-determined body composition. Linear regression analysis was used to assess the relationship between individual biomarkers and outcomes. To reduce multiple hypothesis burden, we used the group biomarkers associated with MD (p adj ≤ 0.05) to assess biomarker level against functional assessments and MRI outcomes. Similarly, we used the paired change in GC responsive markers (p adj ≤ 0.05) against change in outcomes to assess the relationship of pharmacodynamic markers with individual patient outcomes. Patients with missing test values were omitted from the analysis. For each analysis, coefficients and p-values were extracted and FDR hypothesis correction was applied.

## RESULTS

### Proteins differentially present in serum from adults with MD

The WSiMD cohort was previously described ^11^, and included 19 individuals with LGMD and one adult with BMD. Of participants with LGMD, 9 participants had subtype 2A (*CAPN3*), 3 participants had subtype 2B (*DYSF*), 2 participants had subtype 2I (*FKRP*), 1 participant had type 2J (*TTN*), and 2 participants had type 2L (*ANO5*). The participant with BMD had an exon 45-47 deletion in *DMD*. The ages of participants ranged between 18 and 60, with a mean age of 35. Thirteen participants were male, and 7 were female. Six participants were non-ambulatory, 9 were ambulatory, and 5 were transitioning from ambulatory to non-ambulatory status. The control group was age- and sex-matched (**Table 1**), and was not exposed to steroid use, and therefore enabled baseline comparisons between the WSiMD cohort and individuals without muscular dystrophy.

**Table 1.** WSiMD and Control Cohort Demographics.

|  | **LGMD** |  | **CONTROL** |  |
| --- | --- | --- | --- | --- |
|  | n | % | n | % |
| **Sex at birth** |  |  |  |  |
| male | 13 | 68.4 | 10 | 62.5 |
| female | 6 | 31.6 | 6 | 37.5 |
| **Age** |  |  |  |  |
| Median (Yr) | 37 |  | 37 |  |
| range | 18-60 |  | 25-66 |  |
| **LGMD subtype (gene)** |  |  |  |  |
| 2A-CAPN3 | 9 | 47.4 | 0 | 0 |
| 2B-DYSF | 3 | 15.8 | 0 | 0 |
| 2C-SGCG | 1 | 5.3 | 0 | 0 |
| 2I-FKRP | 2 | 10.5 | 0 | 0 |
| 2J-TTN | 1 | 5.3 | 0 | 0 |
| 2L-ANO5 | 2 | 10.5 | 0 | 0 |
| BMD-DMD | 1 | 5.3 | 0 | 0 |

We used an aptamer-based method to measure over 6000 proteins in the serum of WSiMD participants. We analyzed MD-responsive biomarkers using a linear mixed effects model by comparing aptamer profiles from baseline WSiMD participants with age- and sex-matched healthy controls. This comparison identified 507 proteins with an adjusted p value < 0.05 and an absolute log fold change > 0.5. Of these, 109 (22%) were upregulated and 398 (78%) were downregulated in WSiMD participants at baseline compared to healthy controls (**Fig 1A**). These proteins upregulated in the serum were mainly structural sarcomeric proteins, markers involved in skeletal muscle function and differentiation, and metabolic proteins, consistent with leak of muscle proteins into the serum reflecting ongoing muscle injury that is common across many MD subtypes (**Fig 1B**). The sarcomeric proteins ACTN2, MYL3, TNNI2, MYOM2, MYOM3, CFL2, TNNT3, TTN, TPM3, CFL2, PDLIM3, PDLIM5, ANKRD2, TNNT2, TNNI2 were the most upregulated family of biomarkers, by representation and degree of upregulation. The clinically measured protein CKM, along with CA3, were also in the top 20 proteins most highly associated with disease. Constant muscle turnover and regeneration is a hallmark of muscular dystrophies, and consistent with this, we identified upregulation of several proteins implicated in muscle differentiation and repair including KLHL41, CACNB4, CFL2, PDLIM5, and TRIM72. Additional upregulated serum proteins included long chain fatty acid binding proteins FABP3 and FABP4, and FABP3, and these markers have been previously associated with muscular dystrophy ^12,17,22–24^.

**Figure 1.**
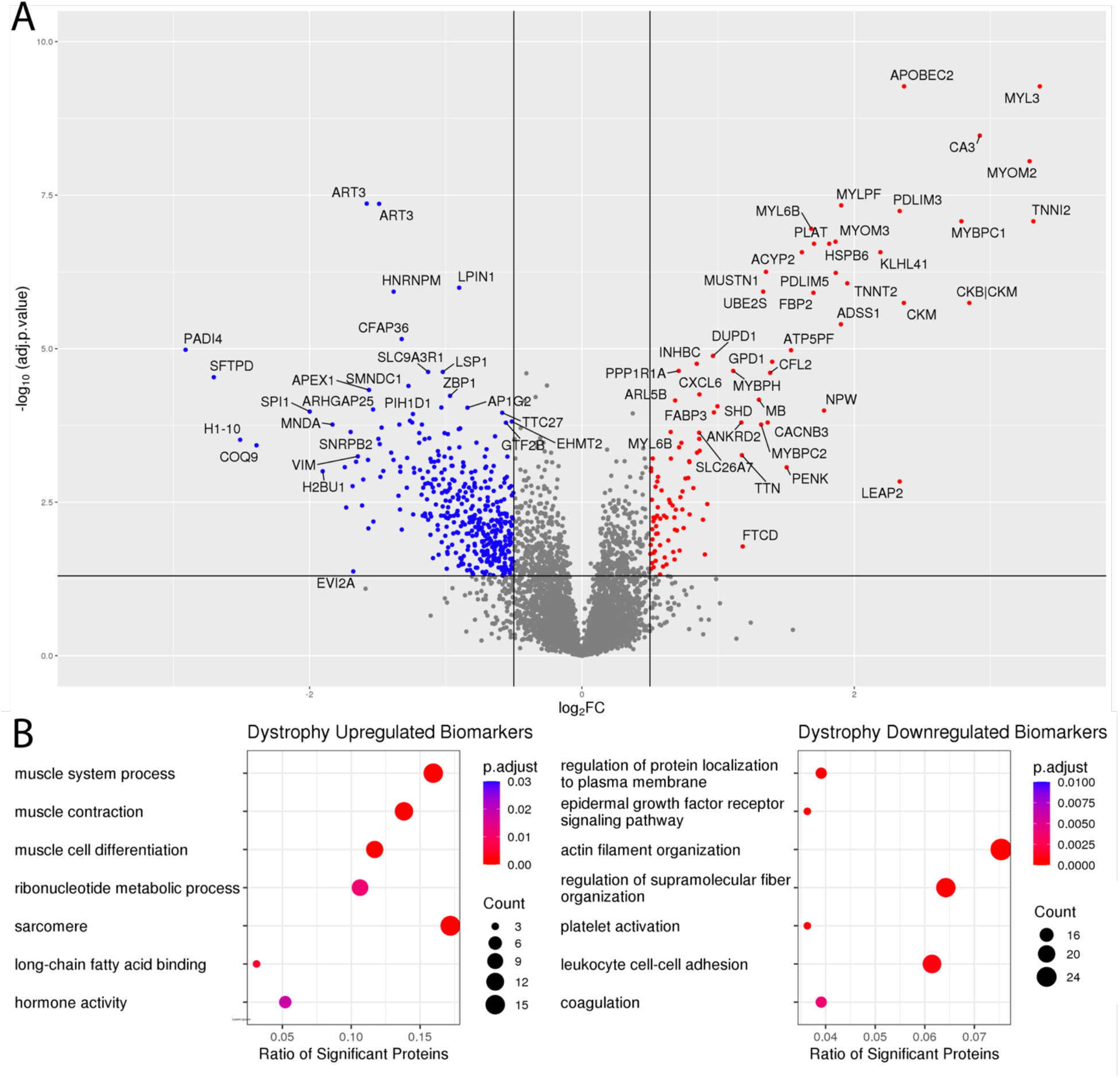
MD Associated Biomarkers. **A)** Volcano plot of aptamer profiling from WSiMD participants compared to age/sex matched controls compared using a linear mixed effects model. Benjamini-Hochberg corrected p-value < 0.05 and abs(log2FC) > 0.5 were used as thresholds for significance. **B)** Gene Ontology terms were used to cluster the upregulated (upper panel) and downregulated (lower panel) groupings of protein biomarkers. Upregulated proteins were largely proteins important for muscle structure, function, and development. Downregulated proteins included those implicated in cellular adhesion, membrane fusion, actin organization, growth factor signaling and coagulation.

Proteins downregulated in the serum of the WSiMD cohort were those implicated in cellular adhesion, membrane fusion, actin organization, growth factor signaling and coagulation (**Fig 1B**). The membrane fusion proteins STX4, and STX8 and the vesicle trafficking/fusion protein VTI1B were significantly downregulated. RHOQ and EZR, which are both involved in the organization and stabilization of the actin cytoskeleton, were decreased, as was the Src family kinases downstream of Fc-receptor signaling LYN, FGR, and FYN. VAV1 and VAV3, both involved in regulation of intracellular calcium levels were reduced in serum from WSiMD participants compared to healthy controls. The platform lacks an aptamer for VAV2, whose catalytic efficiency has been positively associated with muscle mass ^25^. ANXA1, a membrane stabilizing and immunosuppressive biomarker, was also reduced in WSiMD participants compared to healthy controls.

### Prednisone-responsive biomarkers in the WSiMD cohort

**Fig 2** shows a hierarchical clustered heatmap assessing both disease and treatment status across both the WSiMD and control cohorts. When compared in this manner, differences associated with disease-status were more evident than the effect of steroid treatment, yet this analysis also separated patterns by treatment status, distinguishing between prednisone naïve and treated participants. To assess the effect of six months of intermittent prednisone treatment, we evaluated the baseline and study endpoint serum aptamer profiles in the WSiMD participants who received six months of intermittent prednisone. We found that glucocorticoid dosing significantly upregulated 24 proteins and downregulated 132 proteins with an adjusted p-value < 0.05, and an absolute value logFC > 0.2 (**Fig 3A**). Twenty-four proteins had adjusted p-values < 0.01. Consistent with the known effect of glucocorticoids on the immune system, most of downregulated ontology terms associated broadly with immune system function and additionally we observed muscle structural constituents decreased (**Fig 3B**). The Tumor Necrosis Factor α (TNFα) and interferon gamma (IFN-γ) pathways are both inflammatory pathways activated in response to muscle damage and are implicated in the progression of muscular dystrophies ^26,27^. Intermittent prednisone dosing reduced the IFN-γ-responsive proteins WNT5A, IL12B, and the chemoattractant cytokines CCL21 and CCL22. WNT5A and IL12B are known to be responsive to TNFα ^28,29^. C1QTNF4, CD33, and LILRA4 were also downregulated in this group. Overall, these findings indicate that this relatively low, once-weekly dose of prednisone modulated inflammatory pathways in the WSiMD cohort.

**Figure 2.**
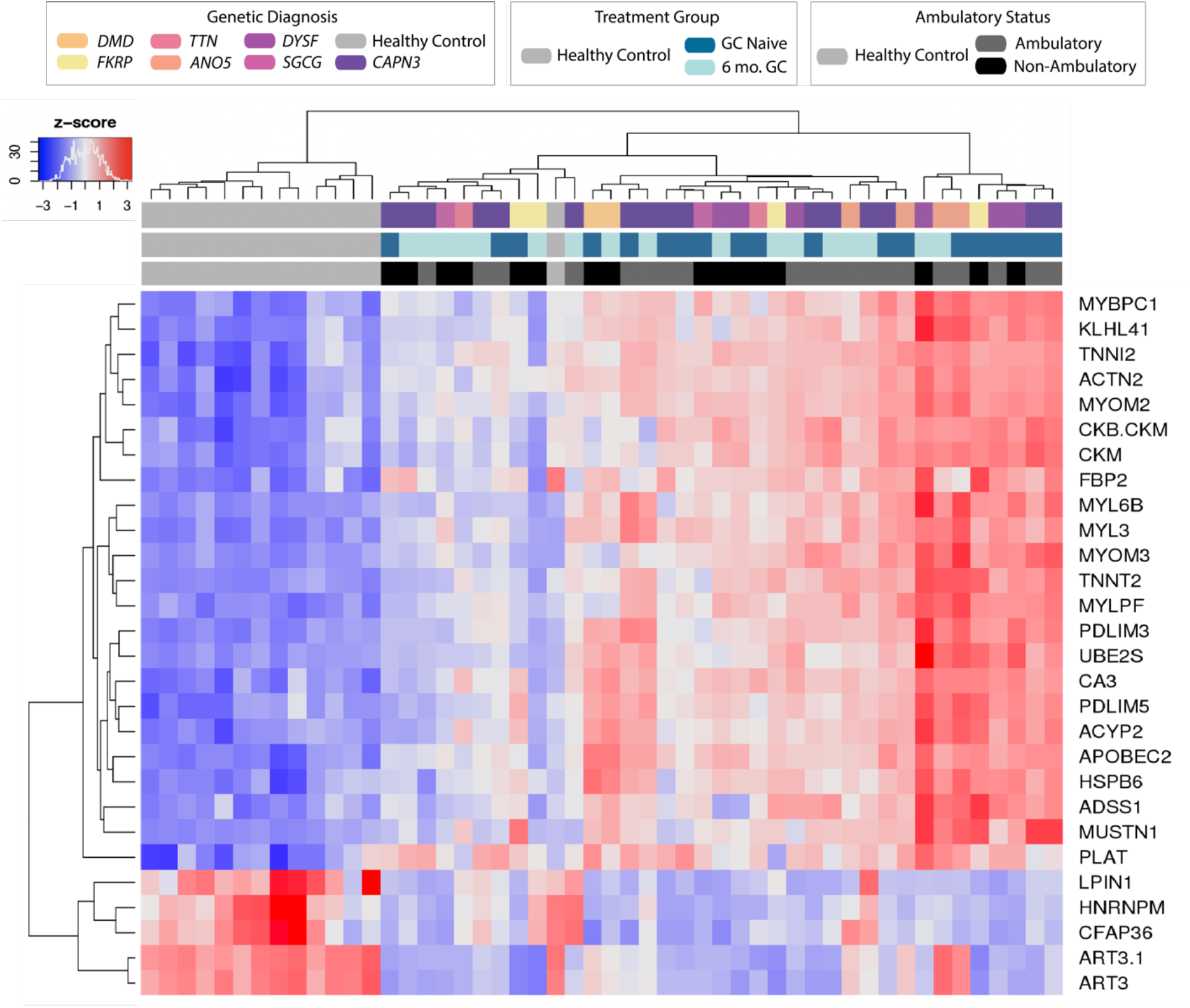
Serum protein markers clustered by disease status and treatment group. Heatmap depicting the behavior of the most significant (p < 0.00001) biomarkers associated with LGMD.

**Figure 3.**
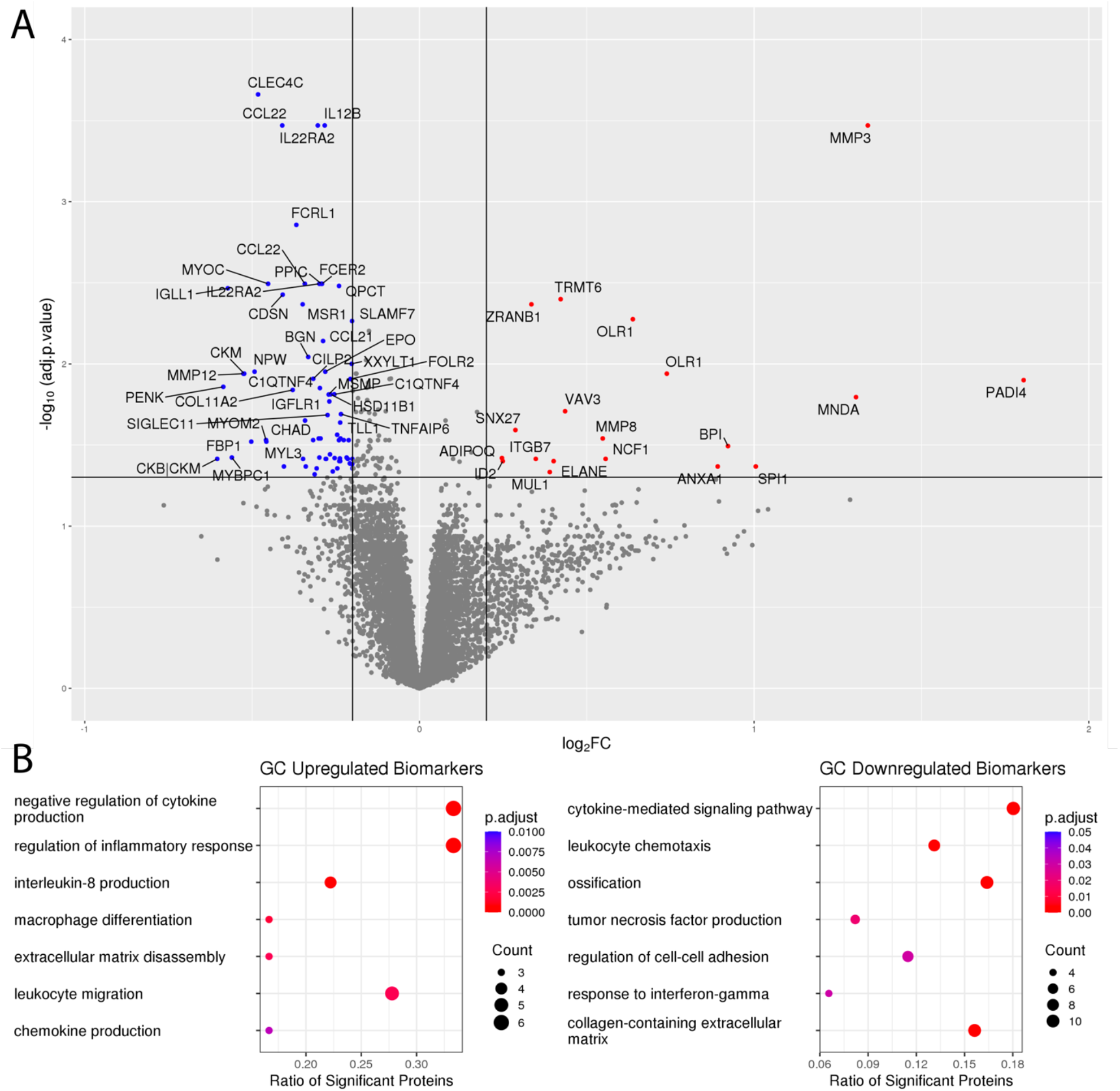
Protein biomarkers in WSiMD participants responsive to 6 months of intermittent glucocorticoid (GC) treatment. **A)** Volcano plot depicting results from linear mixed-effects model for prednisone treatment status. Benjamini-Hochberg corrected p-value < 0.05 and abs(log2FC) > 0.2 were used as thresholds for significance. **B)** Gene Ontology groups of GC up- and down-regulated serum proteins. Upregulated serum proteins are implicated in extracellular matrix disassembly and immune/inflammatory function. Downregulated serum proteins were largely extracellular matrix proteins, proteins involved in muscle structure and function, growth factor signaling, and immune/inflammatory cell adhesion and chemotaxis.

When evaluating upregulated proteins by glucocorticoid treatment, multiple proteases were seen, including ELANE, MMP3, and MMP8 (**Fig 3A**). These components of the “extracellular matrix disassembly” ontology grouping may participate in matrix turnover during regeneration, growth and repair (**Fig 3B**). Prednisone treatment was associated with an increase in transcriptional regulators like ID2, MNDA, SPI1, BPI. Other proteins of interest that were upregulated included ADIPOQ, ANXA1 and MUL1, which are implicated in inflammatory responses and protein turnover, which may be specifically important for muscle growth.

### The effect of prednisone on muscular dystrophy-linked proteins

Since more proteins were differentially present based on disease status, rather than response to prednisone, we assessed which disease-associated ontology terms were shifted towards normalization by exposure to prednisone. This analysis included terms upregulated in muscular dystrophy that were downregulated by prednisone treatment and vice versa (**Table 2**).

**Table 2.**
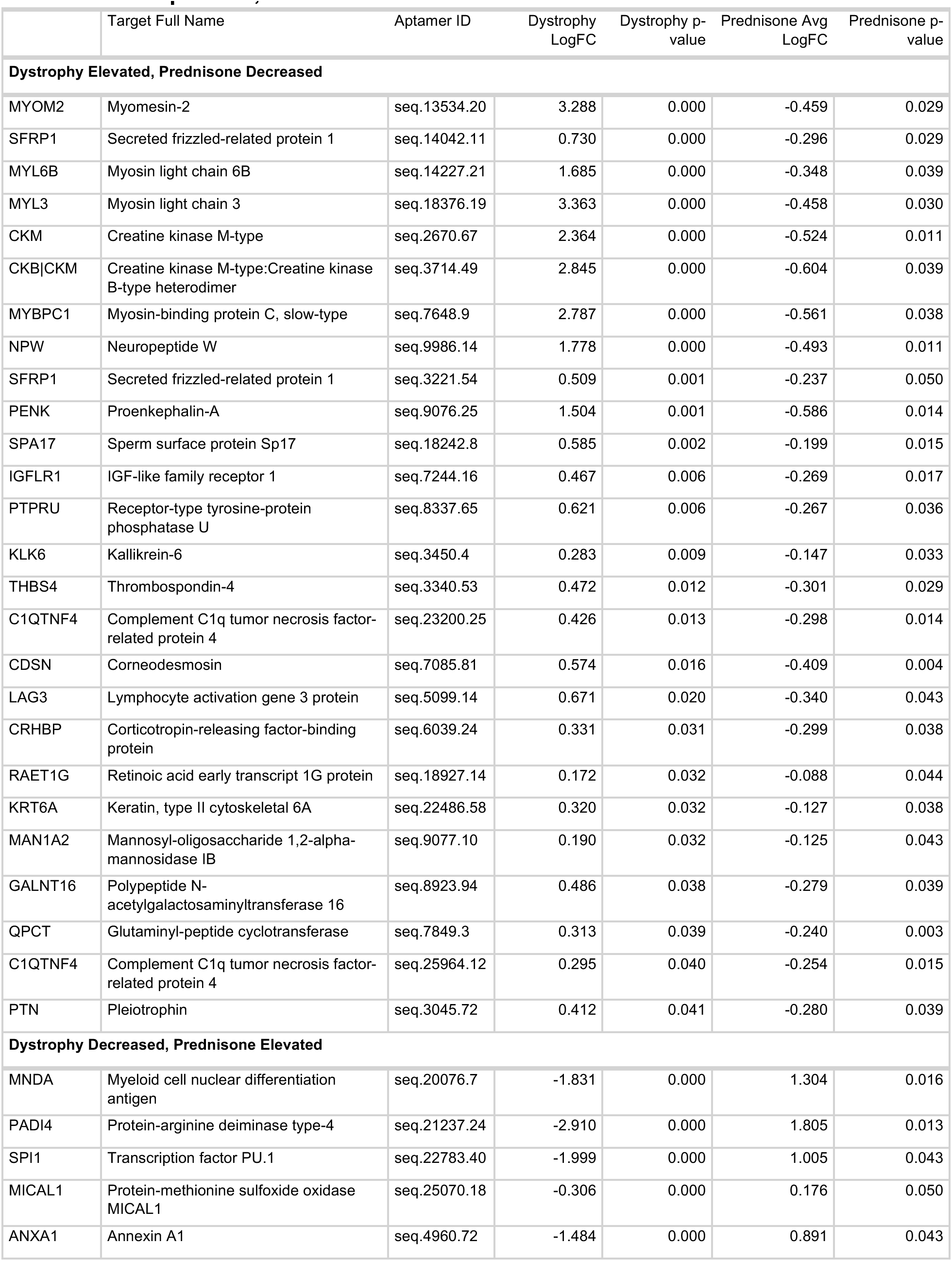
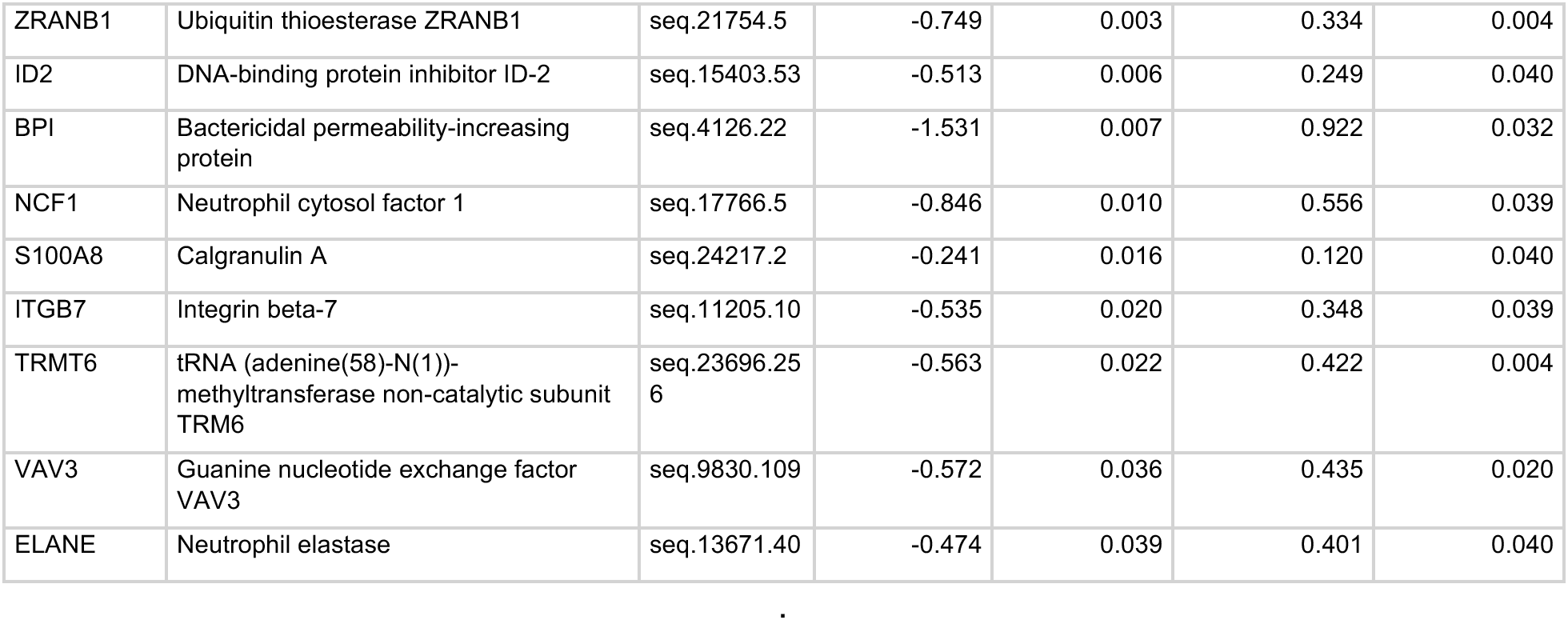
GC responsive, MD-associated biomarkers.

|  | Target Full Name | Aptamer ID | Dystrophy LogFC | Dystrophy p-value | Prednisone Avg LogFC | Prednisone p-value |
| --- | --- | --- | --- | --- | --- | --- |
| **Dystrophy Elevated, Prednisone Decreased** |  |  |  |  |  |  |
| MYOM2 | Myomesin-2 | seq.13534.20 | 3.288 | 0.000 | -0.459 | 0.029 |
| SFRP1 | Secreted frizzled-related protein 1 | seq.14042.11 | 0.730 | 0.000 | -0.296 | 0.029 |
| MYL6B | Myosin light chain 6B | seq.14227.21 | 1.685 | 0.000 | -0.348 | 0.039 |
| MYL3 | Myosin light chain 3 | seq.18376.19 | 3.363 | 0.000 | -0.458 | 0.030 |
| CKM | Creatine kinase M-type | seq.2670.67 | 2.364 | 0.000 | -0.524 | 0.011 |
| CKB\|CKM | Creatine kinase M-type:Creatine kinase B-type heterodimer | seq.3714.49 | 2.845 | 0.000 | -0.604 | 0.039 |
| MYBPC1 | Myosin-binding protein C, slow-type | seq.7648.9 | 2.787 | 0.000 | -0.561 | 0.038 |
| NPW | Neuropeptide W | seq.9986.14 | 1.778 | 0.000 | -0.493 | 0.011 |
| SFRP1 | Secreted frizzled-related protein 1 | seq.3221.54 | 0.509 | 0.001 | -0.237 | 0.050 |
| PENK | Proenkephalin-A | seq.9076.25 | 1.504 | 0.001 | -0.586 | 0.014 |
| SPA17 | Sperm surface protein Sp17 | seq.18242.8 | 0.585 | 0.002 | -0.199 | 0.015 |
| IGFLR1 | IGF-like family receptor 1 | seq.7244.16 | 0.467 | 0.006 | -0.269 | 0.017 |
| PTPRU | Receptor-type tyrosine-protein phosphatase U | seq.8337.65 | 0.621 | 0.006 | -0.267 | 0.036 |
| KLK6 | Kallikrein-6 | seq.3450.4 | 0.283 | 0.009 | -0.147 | 0.033 |
| THBS4 | Thrombospondin-4 | seq.3340.53 | 0.472 | 0.012 | -0.301 | 0.029 |
| C1QTNF4 | Complement C1q tumor necrosis factor-related protein 4 | seq.23200.25 | 0.426 | 0.013 | -0.298 | 0.014 |
| CDSN | Corneodesmosin | seq.7085.81 | 0.574 | 0.016 | -0.409 | 0.004 |
| LAG3 | Lymphocyte activation gene 3 protein | seq.5099.14 | 0.671 | 0.020 | -0.340 | 0.043 |
| CRHBP | Corticotropin-releasing factor-binding protein | seq.6039.24 | 0.331 | 0.031 | -0.299 | 0.038 |
| RAET1G | Retinoic acid early transcript 1G protein | seq.18927.14 | 0.172 | 0.032 | -0.088 | 0.044 |
| KRT6A | Keratin, type II cytoskeletal 6A | seq.22486.58 | 0.320 | 0.032 | -0.127 | 0.038 |
| MAN1A2 | Mannosyl-oligosaccharide 1,2-alpha-mannosidase IB | seq.9077.10 | 0.190 | 0.032 | -0.125 | 0.043 |
| GALNT16 | Polypeptide N-acetylgalactosaminyltransferase 16 | seq.8923.94 | 0.486 | 0.038 | -0.279 | 0.039 |
| QPCT | Glutaminy-peptide cyclotransferase | seq.7849.3 | 0.313 | 0.039 | -0.240 | 0.003 |
| C1QTNF4 | Complement C1q tumor necrosis factor-related protein 4 | seq.25964.12 | 0.295 | 0.040 | -0.254 | 0.015 |
| PTN | Pleiotrophin | seq.3045.72 | 0.412 | 0.041 | -0.280 | 0.039 |
| **Dystrophy Decreased, Prednisone Elevated** |  |  |  |  |  |  |
| MNDA | Myeloid cell nuclear differentiation antigen | seq.20076.7 | -1.831 | 0.000 | 1.304 | 0.016 |
| PADI4 | Protein-arginine deiminase type-4 | seq.21237.24 | -2.910 | 0.000 | 1.805 | 0.013 |
| SPI1 | Transcription factor PU.1 | seq.22783.40 | -1.999 | 0.000 | 1.005 | 0.043 |
| MICAL1 | Protein-methionine sulfoxide oxidase MICAL1 | seq.25070.18 | -0.306 | 0.000 | 0.176 | 0.050 |
| ANXA1 | Annexin A1 | seq.4960.72 | -1.484 | 0.000 | 0.891 | 0.043 |
| ZRANB1 | Ubiquitin thioesterase ZRANB1 | seq.21754.5 | -0.749 | 0.003 | 0.334 | 0.004 |
| ID2 | DNA-binding protein inhibitor ID-2 | seq.15403.53 | -0.513 | 0.006 | 0.249 | 0.040 |
| BPI | Bactericidal permeability-increasing protein | seq.4126.22 | -1.531 | 0.007 | 0.922 | 0.032 |
| NCF1 | Neutrophil cytosol factor 1 | seq.17766.5 | -0.846 | 0.010 | 0.556 | 0.039 |
| S100A8 | Calgranulin A | seq.24217.2 | -0.241 | 0.016 | 0.120 | 0.040 |
| ITGB7 | Integrin beta-7 | seq.11205.10 | -0.535 | 0.020 | 0.348 | 0.039 |
| TRMT6 | tRNA (adenine(58)-N(1))-methyltransferase non-catalytic subunit TRM6 | seq.23696.256 | -0.563 | 0.022 | 0.422 | 0.004 |
| VAV3 | Guanine nucleotide exchange factor VAV3 | seq.9830.109 | -0.572 | 0.036 | 0.435 | 0.020 |
| ELANE | Neutrophil elastase | seq.13671.40 | -0.474 | 0.039 | 0.401 | 0.040 |

Examples of the individual trends in biomarker shifts are shown in **Fig 4**. **Fig 4A** shows the individual responses to MMP3, MMP8, CCL21 and CCL22, demonstrating trends towards normalization in response to prednisone treatment. **Fig 4B** shows muscle proteins upregulated in muscular dystrophy that were downregulated prednisone including CKM, as well as CKB plus CKM heterodimer, TNNI2 and TNNI3, with similar trends evident for MYL3, MYOM2, and MYL6B (**Table 2**). Additionally, there were several proteins involved in receptor ligand activity including the innate immune and TNF-α related proteins, C1QTNF4, PTN, THBS4, and PENK (**Table 2**).

**Figure 4.**
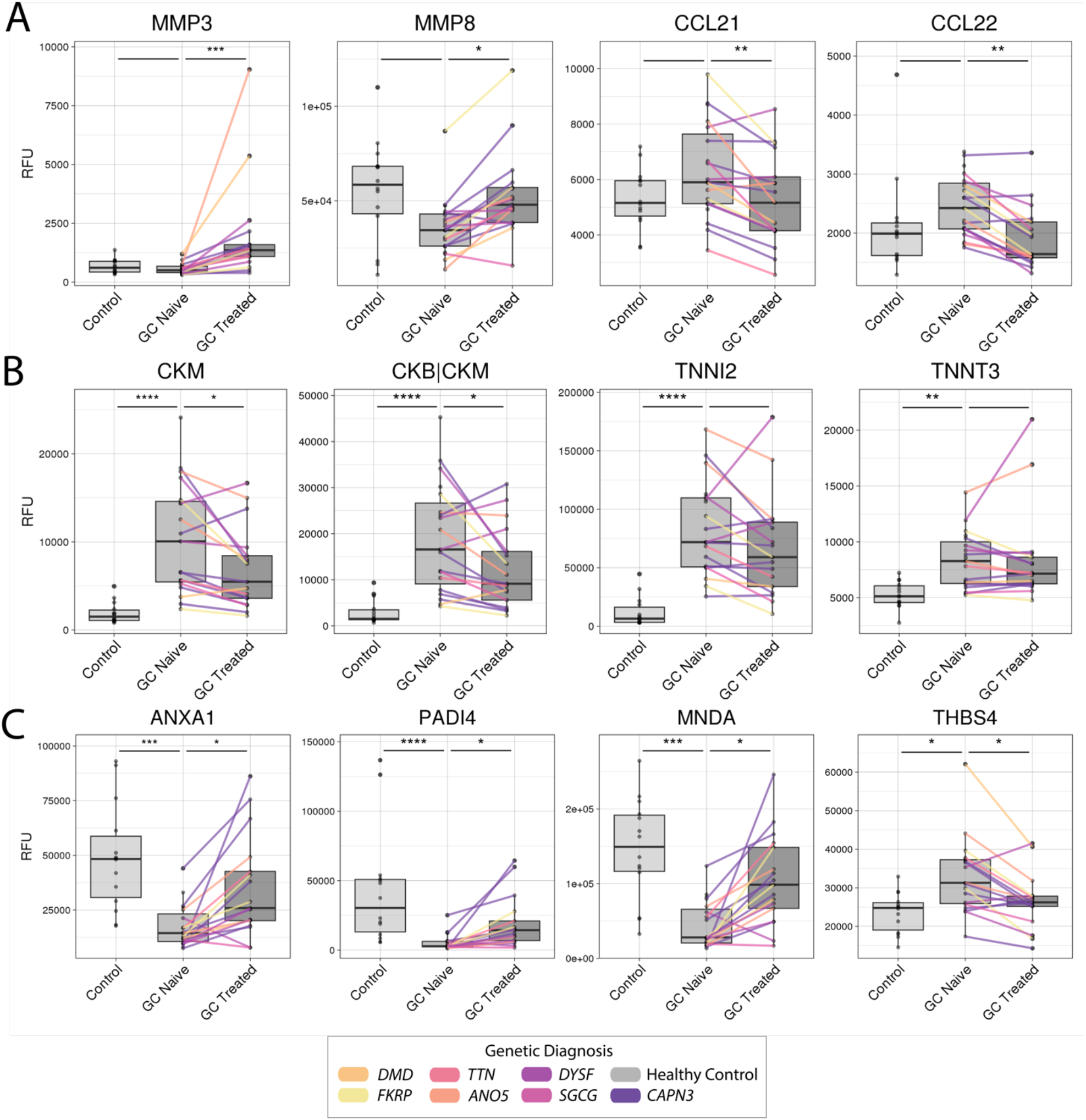
Individual shifts in prednisone- and disease-associated Biomarkers. Boxplots with paired results depicting representative behavior of biomarkers which had a significant response in both LGMD and the paired prednisone comparison. P-values are derived from the linear mixed model, thresholds are (* p < 0.05, ** p < 0.01, *** p < 0.001, **** p < 0.0001). **A)** Glucocorticoid-only responsive marker. **B)** Clinically monitored creatine kinase and troponin markers. **C)** Proteins displaying both dystrophy association and glucocorticoid responsiveness. GC treatment had disease normalizing effects in this group.

Proteins downregulated in MD that were upregulated by prednisone included mainly proteins implicated in immune process. Notably, PADI4, a protein deiminase that can reduce the affinity of chemoattractant ligands was the most downregulated biomarker by logFC in the WSiMD cohort at baseline, and PADI4 was the most upregulated after prednisone treatment (**Fig 3** and **Fig 4B**). Similarly, the membrane stabilizing and immunosuppressive proteins ANXA1, was also upregulated by prednisone treatment (**Fig 3** and **Fig 4B**). SPI1, a master regulator of myeloid cell differentiation was also strongly upregulated by prednisone. Other proteins in this category were ITGB7, ELANE, NCF1, S100A8, BPI, and VAV3 (**Fig 5**).

**Figure 5.**
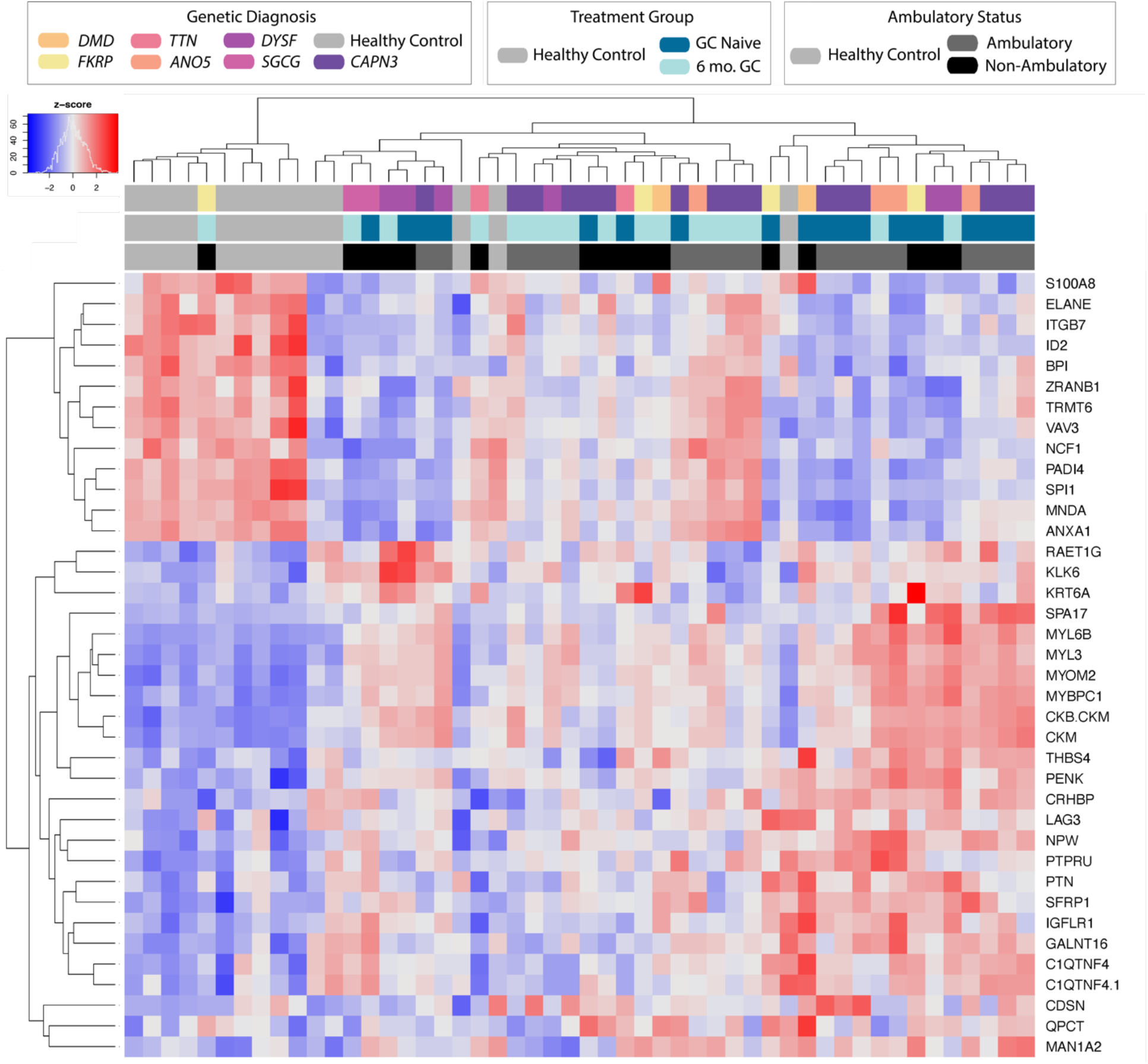
Muscular Dystrophy-associated, GC responsive signature separates control serum profiles from GC treated from GC naïve sera. Heatmap depicting the behavior of muscular dystrophy associated, glucocorticoid responsive serum biomarkers.

Interestingly, prednisone treatment also upregulated ID2, a transcription factor that is in an inhibitor of differentiation. ID2 has been shown to also inhibit MyoD, a myogenic regulatory factor, and this observation coupled with stabilized sarcolemma from an increase in ANXA1 could indicate a lessened need for regeneration with prednisone exposure.

### Comparisons With Previous Studies

We compared the findings from the WSiMD cohort with five prior MD studies that used the similar aptamer platform ^12,17,22,30,31^. These studies included a DMD cohort of young and slightly older patients with DMD, and a study evaluating the response of DMD patients to steroid treatment ^12,30^. It also included a study of FSHD patients ^31^ (**Table 3**). Across all studies comparing results to healthy controls, CKM, CA3, TNNI2 and FABP3 were upregulated. MB and ANP32B were upregulated in the DMD studies comparing to control ^12,17,22^. There was substantial overlap between the list of downregulated proteins in the WSiMD cohort and those upregulated in younger DMD patients ^12^. These proteins included 14-3-3 proteins YWHAB, YWHAH, and YWHAZ, as well as ENO1, XRCC6, EIF4G2, EIF4H, and HNRNPA2B1. This discrepancy might reflect differences in disease etiologies or disease trajectory, or it could reflect inherent variability in these measures. Also of note, in the WSiMD cohort, compared to the younger DMD cohort, there was less immune downregulation and fewer upregulated proteins at baseline. This distinction may reflect the more intense nature of DMD, reflected by its younger age of onset and faster progression to loss of ambulation in DMD compared to the adult MD patients in WSiMD.

**Table 3.**
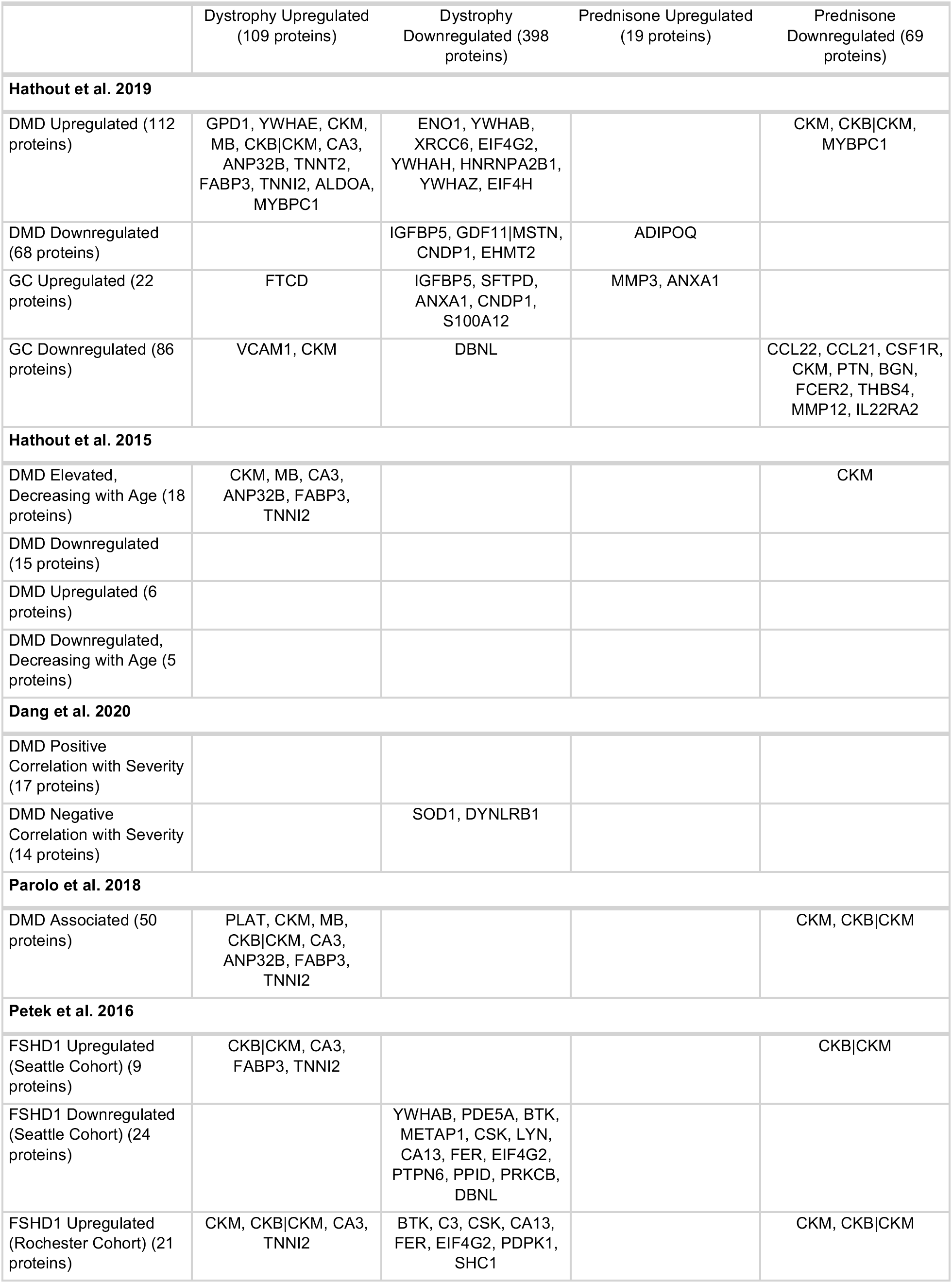
Comparison with previous studies in DMD and FSHD.

The treatment responsive serum proteins in the WSiMD cohort shared some overlap with the glucocorticoid treated DMD group described by ^12^. Notably, 24 weeks of once weekly prednisone in WSiMD yielded a set of biomarkers that had overlap with the Hathout 2019 study, displaying upregulation of MMP3 and membrane-stabilizing and immune-suppressive ANXA1. CCL22, CCL21, CSF1R, CKM, PTN, BGN, FCER2, THBS4, MMP12, and ILL22RA2 were all downregulated in both studies. This “normalizing” effect of once weekly prednisone was similar to higher doses used in the DMD patients and may indicate these biomarkers are especially relevant to mitigating the muscular dystrophy process.

### Integrating MRI, serum profiles and functional status

Despite the small size and genetic and functional heterogeneity of the WSiMD cohort, we assessed for correlations between MR imaging, functional status and serum aptamer profiles. We evaluated fat fraction in the lower extremity studies with focus on the quadriceps muscle group and Triceps surae (**Fig 6A**). We included data for those participants who completed the MRI at study endpoint and for whom there was a sufficient quality image for analysis (n=10 for quadriceps, n=13 for Triceps surae). Overall, most participants showed a small trend towards increasing fat fraction during the open label study. We correlated functional study and body composition as determined by DEXA scanning (**Fig 6B**). We noted a negative correlation between muscle fat fraction and grip strength, as well as North Star Assessment for limb-girdle muscular dystrophies (NSAD). NSAD also showed a high degree of correlation with the other functional assessments. As NSAD was also the most complete dataset, we used it to assess association of disease biomarkers with disease severity by linear regression. We observed a significant association between CKM and NSAD functional status, which reflects what is also seen in DMD, and this same pattern was evident for the muscle marker PGAM2 (**Fig 6C**). We also evaluated the paired changes in GC responsive biomarkers with changes in NSAD. The change in CKM over the 6 months of prednisone treatment suggested that those with greater reduction in CKM had greater gain in functional status, so that change in CKM over this short time frame may reflect improved muscle repair or membrane stability. It is interesting that while higher CKM is associated with better functional status, likely due to increased activity and muscle mass, larger decreases were associated with improved functional outcomes. This observation likely relies on a maintenance or increase in lean mass, as was observed in this cohort. There was also a trend toward improved functional in those participants with a decline in CCL22, a marker previously shown as steroid responsive in DMD ^12^.

**Figure 6.**
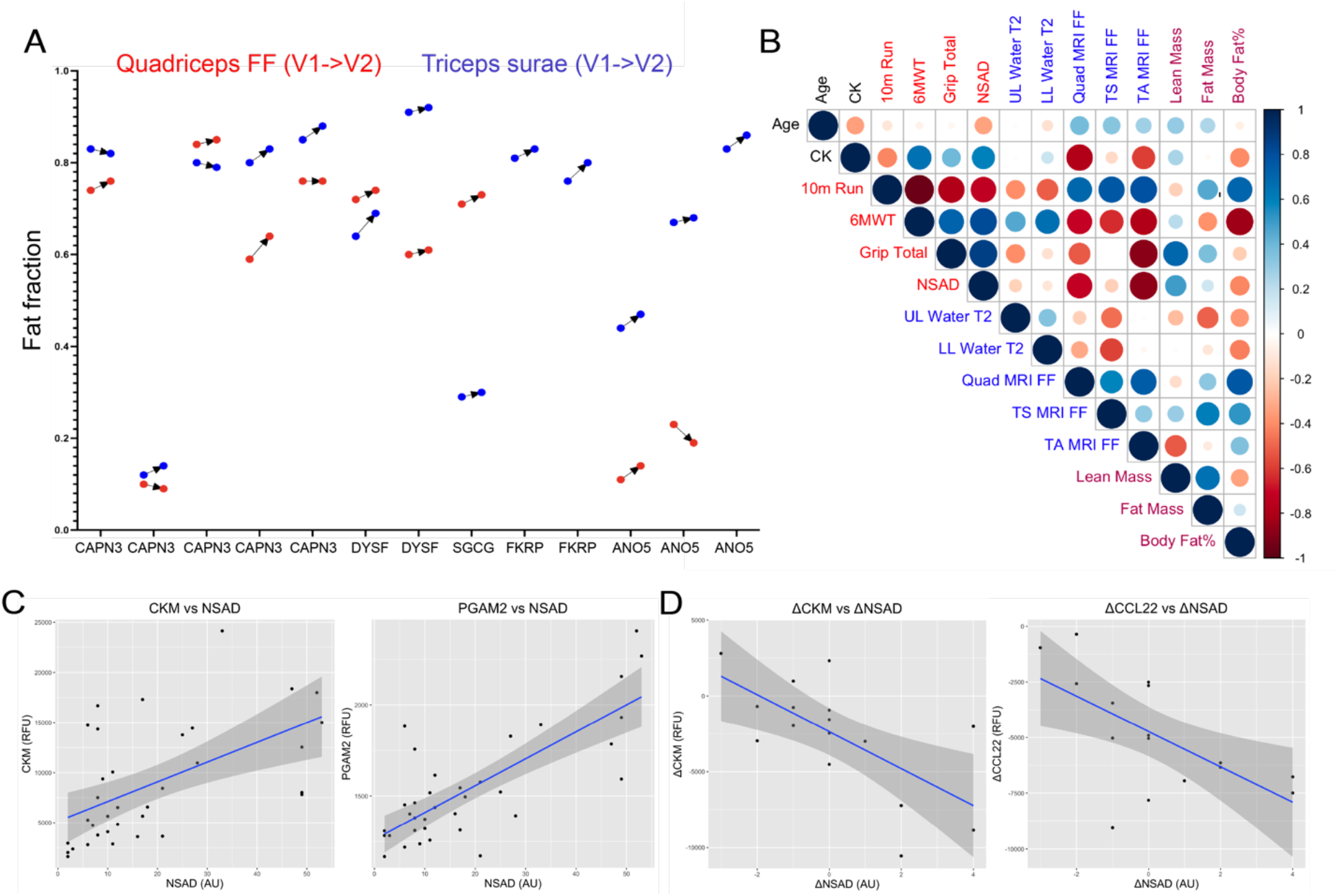
MRI and biomarkers related to functional outcomes. **A)** Muscle fat fraction at study onset and after 6 months of weekly prednisone (V1, visit 1; V2, visit 2) in the quadriceps (RED) and Triceps surae (BLUE) muscle groups. Participants are listed by gene name along the X axis. Data was available from two muscle groups for 10 participants. For the two *FKRP* participants and one *ANO5* participant, data was only available from the Triceps surae (blue). The majority of participants experienced a small increase in muscle fat fraction in the quadriceps and Triceps surae muscle groups. There was not always agreement between the two muscles. **B)** Spearman correlation coefficients between age, functional performance measures, MRI water T2 and muscle fat fraction, and body composition as determined by DEXA scanning. **C)** Example biomarkers showing a significant association (FDR ≤ 0.05) by linear regression with NSAD measure. The muscle protein markers CKM and PGAM2 correlated positively with functional performance as assessed by NSAD. **D)** Broad trends towards CKM and CCL33 reduction with change in NSAD over six months of treatment with once weekly prednisone (p value ≤ 0.05, FDR ≥ 0.05).

## DISCUSSION

### Steroid use in muscular dystrophy

The WSiMD study was conceived to provide pilot information to guide a larger study of steroid use for MD subtypes not typically treated with glucocorticoids. Although DMD patients receive glucocorticoids with demonstrated functional benefit, this treatment comes with significant adverse outcomes, especially over the lifetime of a DMD patient ^32^. These adverse consequences are the commonly observed side effects that accompany long term glucocorticoid use for any condition. Long term adverse effects of obesity, metabolic syndrome/insulin resistance, adrenal insufficiency, cataracts, and osteoporosis are known consequences of years of exogenous glucocorticoid exposure. In DMD, glucocorticoid use typically begins in young children, and prolonged steroid during childhood stunts bone growth, body growth, and puberty. To this end, the novel glucose-sparing vamorolone received approval in the US and EU in 2023 for treating DMD based on its efficacy in boys with DMD ^33,34^. For vamorolone, it is encouraging that bone growth appears less adversely affected in DMD compared to prednisone ^33,34^. The degree to which vamorolone protects against the other complications from prednisone or deflazacort use will be determined as more DMD patients transition to vamorolone for lifetime use.

An alternative strategy of using intermittent steroids has been employed in DMD, and one advantage of this intermittent dosing is less adrenal insufficiency, obesity, and insulin resistance ^6,9^. In a study comparing 10 day on/off with daily glucocorticoids, intermittent steroids had less benefit on functional outcomes ^3^. This study did not examine high dose weekend dosing, which is likely to have distinct effects from the 10 day on/off regimen. While some intermittent glucocorticoid treatment strategies may have less benefit functionally over the shorter term in DMD, the long term outcomes with respect to side effects and how these findings translate to adults with LGMD is simply not known.

For the LGMD subtypes evaluated in this study, especially *CAPN3*, *DYSF*, and *ANO5*, these forms of muscular dystrophy are usually diagnosed in adulthood, and most patients with these subtypes will report having normal muscle function as children. Each of these muscular dystrophy subtypes have elevated serum CK, which if measured in early life, can be used for early diagnosis. Indeed, all the patients participating in the WSiMD trial were adults, and in this population, bone growth and puberty suppression are not clinical concerns as they are in DMD. For lifetime glucocorticoid treatment in LGMD and BMD adults, adverse outcomes of obesity, insulin resistance, hyperlipidemia, and adrenal insufficiency are serious risks that would need to be balanced against any functional benefit.

### Diversity of response to intermittent steroids and using biomarkers to guide steroid dosing

The overall outcomes in this small pilot study were highly variable, which was expected based on inclusion criteria. We noted that a subset of patients appeared to have more benefit in this short duration study, showing improved lean mass ^11^. We did not observe striking shifts in MRS muscle fat fraction, with most participants trending towards a modest increase over the 6 month study. We did observe trending correlations across all measures of muscle function with each other, and to a small degree with upper limb fat fraction and muscle function. A longer duration and placebo controlled glucocorticoid study is needed to determine muscle fat fraction trajectory over time and whether it correlates with muscle function.

With respect to serum biomarkers, we did observe a number of proteins consistent with a response to prednisone treatment, even at this low dose and intermittent exposure, and these markers included proteins both increased and decreased by glucocorticoids. ADIPOQ was increased by glucocorticoids, consistent with prior reports in DMD steroid use ^12^. We recently showed that intermittent prednisone mitigated diet-induced obesity in mice, and the effect of intermittent prednisone required the *Adipoq* gene to achieve this result. In this model, intermittent steroid exposure boosts ADIPOQ production from adipocytes, and correspondingly increases the adiponectin receptor (*AdipoR1*) in skeletal muscle ^35^. Once activated, pathways downstream of the receptor activate CaMKK2 and AMPK to enhance skeletal muscle uptake of nutrients and ATP production. We previously found that mild exercise in *mdx* mice also activated ADIPOQ pathways, suggesting activity in combination with prednisone may be even more effective ^36^.

The serum proteins downregulated by weekly steroid exposure in WSiMD participants that were observed as being steroid responsive in DMD included several proteins implicated broadly in immune function like CCL22, CCL21, CSF1R, PTN, FCER2, and IL22RA2. CCL22 is a chemokine produced by macrophages, dendritic cells and T cells, and is a steroid responsive chemokine that can be used to predict steroid resistance in other settings ^37,38^. Moving forward, it may be possible to design serum markers to more carefully tailor steroid regimens the optimize efficacy and minimize adverse consequences.

### Study limitations

This study is limited by the small sample size, open label and pilot nature of WSiMD, and heterogenous nature of the cohort being studied with respect to primary gene mutation and stage of clinical progression. Despite these limitations, this data can help guide larger and more informative studies to use glucocorticoids to treat muscular dystrophy beyond DMD.

## AUTHOR CONTRIBUTIONS

AZ collected data and samples. AW performed the biomarker analysis. AB performed the MRI analysis. AB, GW, KV analyzed the images. AW, AZ, AD, and EM conceived the studies, analyzed the data, and wrote the manuscript.

## Data Availability

Data that support the findings in this study are included in this article or are available upon
request from the corresponding author and will be shared in manner to protect the privacy of the
participants.

## ACKNOWLEDGEMENTS

Supported by the Kurt+Peter Foundation, National Institutes of Health AR052646, NS047726, NS127383, and HL061322 and Parent Project Muscular Dystrophy. We thank Dr. Todd Parrish and the Center for Translational Imaging at Northwestern.

## Abbreviations

LGMD: Limb Girdle Muscular Dystrophy
MD: muscular dystrophy

